# Pre operative fitness score accurately predicts uneventful post operative course in gastrointestinal and hepatobiliary surgery

**DOI:** 10.1101/2020.04.14.20057612

**Authors:** Bhavin Vasavada, Hardik patel

## Abstract

**Background:** Aim of our study was to analyse if we can accurately predict uneventful post operative course pre operatively in gastrointestinal and HPB surgery patients.

**Material and Methods:** We retrospectively evaluated patients who have undergone gastrointestinal and hepatobiliary surgery at our institute in last 3 years and analysed 90 days mortality and morbidity rates among these patients. We described any 90 day morbidity and mortality as an “event”. We performed univariate and multivariate analyses for factors predicting an “event”. Then based on pre operative factors that predicted an “event” we formulated a score. Statistical analysis was done using SPSS version 23.

**Results:** Total 264 patient operated for gastrointestinal and HPB surgeries between April 2016 to may 2019 were evaluated .Total 45 (17%) events occurred. On univariate analysis CDC grade, ASA score,Operative time,Blood products used, emergency surgeries and open surgeries predicted an event. We developed score based on pre operative factors like ASA score, CDC grade of surgery,open surgery and emergency surgeries included in the score. We proposed score grater than 2 was associated with 90 day event. This score had sensitivity of 77.78%, specificity of 81.65%. low positive predictive value of 46.67% but very high negative predictive value of 94.68%. AUROC showed AUROC of 0.797 (p < 0.0001, 95% confidence interval 0.721-0.874). Pre operative fitness score, Open Surgery and operative time independently predicted an “event” on multivariate analysis. (p =0.003 and 0.026 respectively.

**Conclusions:** Pre operative fitness score accurately predicts uneventful post operative course in gastrointestinal and hepatobiliary surgery.

## Background

Gastrointestinal, hpatobiliary and pancreatic surgeries are on the most commonly performed procedure world wide, However they are one of the most complex procedures also. 90 day Morbidity and mortality after gastrointestinal and hepatobiliary surgeries remains very high despite technical advancements.

[1,2,3,4].

These high mortalities and morbidities force us to predict pre operatively which patients will have higher chances of morbidity and mortality to accurately assess prognosis or to predict safety of the procedure and to evaluate how aggressive we can go to cure the disease and also to make patient understand the risk involved and help them to take informed decision. □

### AIMs of the study

Aims of this study was to evaluate our data for 90 days morbidity and mortalities and to evaluate factors responsible for that. Based on our statistical evaluation we tried to formulate a score based in pre operative evaluation to predict 90 days morbidity and mortality on a particular patient pre operatively or to predict uneventful 90 day post operative course.

## Material and Methods

We retrospectively evaluated patients who have undergone gastrointestinal and hepatobiliary surgeries at our institute in last 3 year and analysed 90 days mortality and morbidity among these patients. We defined morbidity as any grade 3 grade 4 clavien dindo classification. [5]

We described any 90 day morbidity and mortality as an “event”. We performed univariate and multivariate analyses for factors predicting an “event”. Then based on pre operative factors that predicted an “event” we formulated a score and then evaluated sensitivity, specificity, positive predictive and negative predictive value of that score and also evaluated ROC curve and again performed univariate and multivariate analysis of an “event” to check weather the score developed by us independently predicted the outcome or not. Statistical analysis was done using SPSS version 23. Chi square test was done for categorical values, Mann Whitney U test for continuous variables. Multivariate analysis was done using binary logistic regression method. We also evaluated kaplan meier survival curve with log rank analysis for 90 days event free survival.

## RESULTS

Total 264 patient operated for gastrointestinal and HPB surgeries between April 2016 to may 2019 were evaluated retrospectively for any 90 days morbidity and mortality “event”. Total 45 (17%) events occurred. Over all 90 days morbidity and mortality rates were 11.7% and 8.7% respectively.

On univariate analysis CDC grade, ASA score,Operative time,Blood products used, emergency surgeries and open surgeries predicted an event. [Table 1]

**Table 1:** Univariate analysis between “EVENT” and control (uneventful post operative course) group

| Factors | Control group (n= 219) | Event group (n=45) | P value |
| --- | --- | --- | --- |
| Age (mean /SD) | 51.19 (11.75) | 51.64(11.48) | 0.893 |
| CDC Grade of surgery.(mean/SD) | 2.48 (0.655) | 3.15 (0.551) | <0.0001 |
| Blood prodcuts used (mean/SD) | 0.83 (3.55) | 2.45 (5.82) | <0.0001 |
| ASA (mean/SD) | 2.3 9 (0.54) | 2.94 (0.73) | <0.0001 |
| Operative time (minutes) (mean/SD) | 112.75 (109.71) | 184.25 (148.14) | <0.0001 |
| Hospital stay (days) | 3.11 ( 1.92) | 4.19 (2.11) | <0.0001 |
| Open surgery (n) | 104 | 43 | <0.0001 |
| Emmergency Surgery (n) | 32 | 15 | 0.003 |
| Pre operative fitness score >2 (n) | 40 | 35 | < 0.0001 |

### Pre Operative Fitness Score

We developed score based on pre operative factors like ASA score grater than 2, CDC grade of surgery grater than 2,open surgery and emergency surgeries included in the score. Each variable was given 1 point.We proposed score grater than 2 was associated with 90 day event. This score had sensitivity of 77.78%, specificity of 81.65%. low positive predictive value of 46.67% but very high negative predictive value of 94.68%. [Table 2]

**Table 2:** Pre operative fitness score: ASA (Americal society of anesthesia), CDC (centre of disease control). 3 out of 4 factors of score grater than 2 was associated with aderse surgical outcomes or score less then 2 predicted uneventful post operative course. Sensitivity -77.78%, specificity 81.65%, positive predictive value 46.67% and negative predictive value of 94.68%.

| Factors | Points |
| --- | --- |
| Open Surgery | 1 |
| Emmergency Surgery | 1 |
| ASA grade >2 | 1 |
| CDC grade of surgery | 1 |

ROC curve showed AUROC of 0.797 (p < 0.0001, 95% confidence interval 0.721-0.874). [Figure 1]

**Figure 1.**
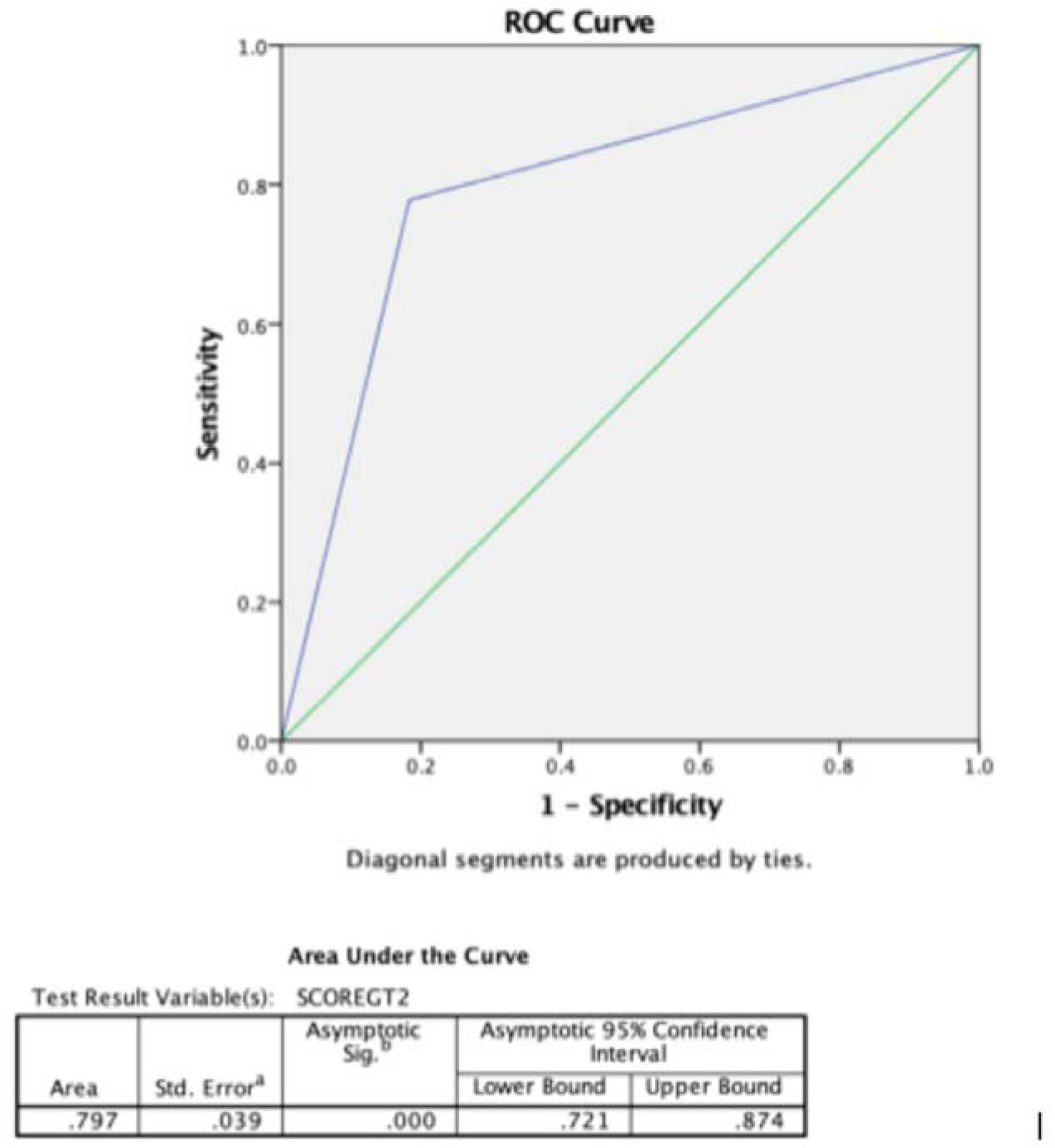
On ROC curve Pre operative fitness score was significantly associated with post operative out comes. (p< 0.0001) with AUROC of 0.797.

### Univariate and multivariate analysis after Including Pre Operative Fitness score

On univariate analysis after including Pre Operative Fitness score open surgery, pre operative fitness score grater than 2, Emmergency Surgery, More operative time,Higher ASA grade and Higher CDC grade of surgery predicted an “event”

Pre operative fitness score and Open surgery independently predicted an “event” on multi-variant analysis. (p =0.003 and 0.026) respectively. [Table 3.]

**Table 3:** Multivariate analysis showed Score greater than 2 and Open surgery independently predicted Surgical outcomes

|  | B | S.E. | Wald | df | Sig. | Exp(B) | 95% C.I. for EXP(B) |  |
| --- | --- | --- | --- | --- | --- | --- | --- | --- |
|  |  |  |  |  |  |  | Lower | Upper |
| Step 1<sup>a</sup> bloodprodcuts | .156 | .158 | .979 | 1 | .322 | 1.169 | .858 | 1.591 |
| operativetime | .005 | .003 | 3.593 | 1 | .058 | 1.005 | 1.000 | 1.011 |
| SCOREGT2(1) | -1.866 | .637 | 8.571 | 1 | .003 | .155 | .044 | .540 |
| openlap(1) | -1.822 | .818 | 4.961 | 1 | .026 | .162 | .033 | .804 |
| EMMERGENC Y(1) | .207 | .584 | .126 | 1 | .723 | 1.230 | .392 | 3.865 |
| ASA | -.179 | .413 | .187 | 1 | .666 | .836 | .372 | 1.881 |
| gradeofsurgery | .187 | .478 | .153 | 1 | .696 | 1.206 | .472 | 3.079 |
| Constant | -1.230 | 2.013 | .373 | 1 | .541 | .292 |  |  |

a. Variable(s) entered on step 1: bloodprodcuts, operativetime, SCOREGT2, openlap, EMMERGENCY, ASA, gradeofsurgery.

On Kaplan Meier Survival analysis score less than 2 was associated with significant higher 90 days “event” free survival rates. (p < 0.0001) [Figure 2].

**Figure 2:**
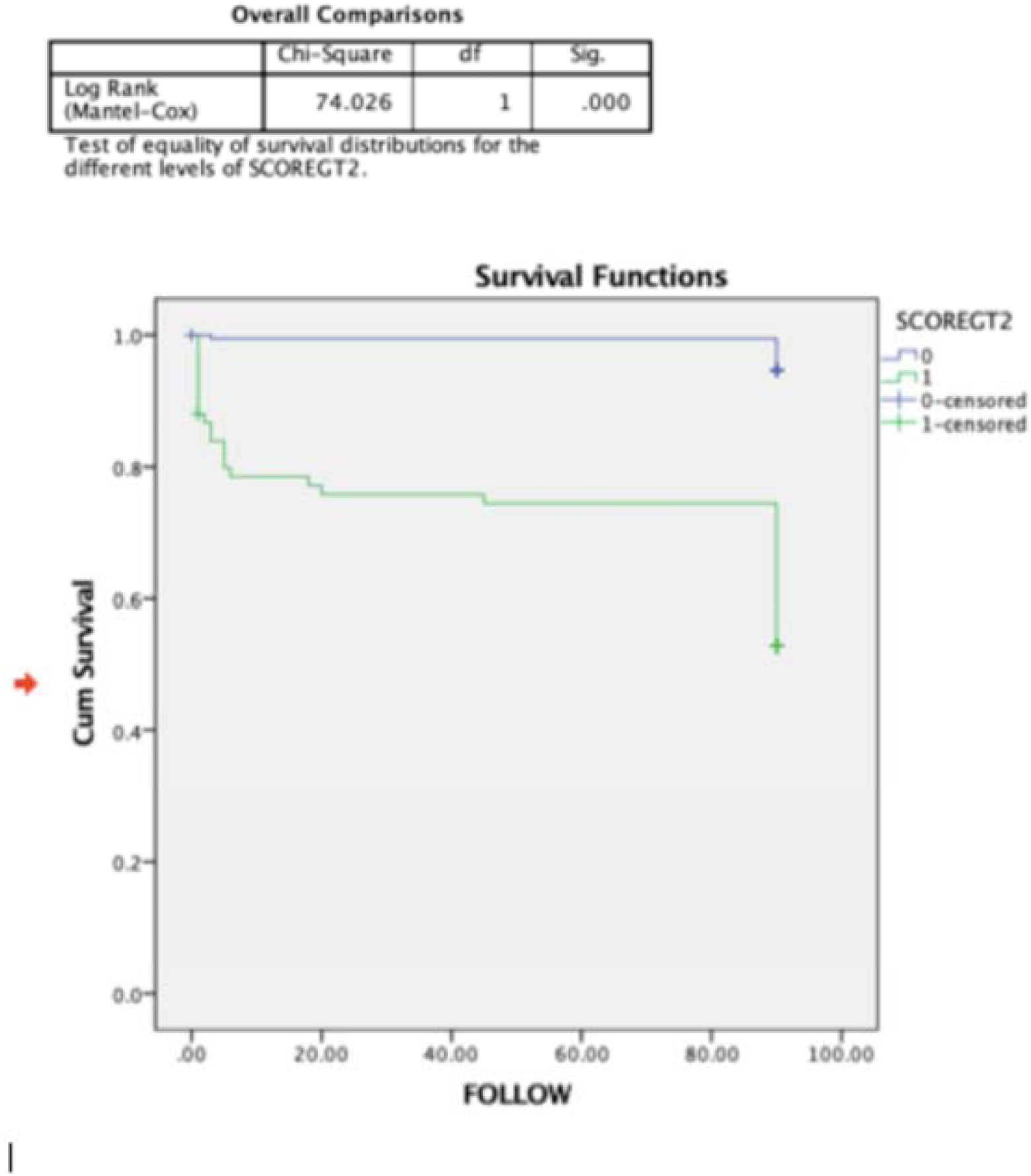
90 days event Free survival was significantly more in pre operative fitness score less than or equal to 2. (p<0.0001)

## Discussion

Surgeons are always worried about out comes and safe surgery is always their goal. Pre operative prediction of surgical outcomes is like holy grail of surgery. In this manuscript we have tried to evaluate our 90 days morbidity and mortality and factors responsible for it and tried to assess pre operative prediction of patients who are likely to develop complications or patients who are likely to have favourable outcomes and thus to accurately assess risk benefit ratio of any procedure. CDC graded surgery and wound according to complexity of diseases and shown that wound complications and surgical site infection rates are higher with higher grades of surgery.[6]

Anaesthetists commonly use American society of Anaesthesia grading [ASA] of physiologic status to evaluate anaesthetic related complication. The American Society of Anaesthesiologists Physical Status (ASA PS) classification system was first introduced in 1941.[7].Various studies have shown that ASA grading is useful in predicting Surgical outcomes. [8,9,10]. We evaluated various factors for prediction of post operative “event” as described above. On univariate analysis we found that ASA score,CDC grade of surgery, more blood products used, increased operative time, open surgeries compared to laparoscopic surgeries, emergency surgeries were associated with increased morbidity and mortality in gastrointestinal and hepato-pancreatico biliary surgery.Various authors have also found similar factors associated with post operative morbidity and mortality.[1,2.3.4].

Various authors have tried to predict post operative outcomes by preoperative factors in various major surgeries [11,12,13,14] but to our knowledge vary few have tried to evaluate a score in major gastrointestinal and hepatobiliary surgeries. From univariate analysis we selected all the preoperative factors affecting post operative outcome and gave each factor one point and evaluated score greater than 2 or if 3 out of four factors positive i.e. ASA score grater than 2, emergency surgery, CDC grade of surgery greater than 2 and open surgery for sensitivity, specificity, positive and negative predictive value and it showed sensitivity and specificity of 77.78% and 81.65% respectively and positive and negative predictive values of 44.67% and 94.68%. Low positive predictive value shows that high score do not preclude the surgery at the same time high negative predictive value suggested that if score is less than 3 it very accurately predicted safe out come in post operative period and it helps us to go little more aggressive in certain diseases. We again performed univariate and multivariate analysis after the score included along with all the factors included in the score and at the end of multivariate analysis it shows that pre operative fitness score independently predicted outcomes after surgery.

AUROC of the score also showed that score significantly associated with an “event” with p values <0.0001 and AUROC of 0.797. These results with high negative predictive value shows that this score can help us to prognostify patients regarding risk and benefit of the surgery and we can plan our risk benefit ratios according to that.

Being a retrospective evaluation this study has inherent bias associated with retrospective analysis. Also multicenter analysis with large volume studies can help to validate these findings further. In conclusion pre operative fitness score predicts post operative outcomes in gastrointestinal and hepato pancreaticobiliary surgery and more accurately predicts uneventful surgery if pre operative fitness score less than 3.

## Data Availability

data will be made available on demand.

